# The duration of lithium use and biological ageing: telomere length, frailty, metabolomic age and all-cause mortality

**DOI:** 10.1101/2023.12.18.23300105

**Authors:** Julian Mutz, Win Lee Edwin Wong, Timothy R. Powell, Allan H. Young, Gavin S. Dawe, Cathryn M. Lewis

## Abstract

**Background:** Lithium is an established first-line treatment for bipolar disorder. Beyond its therapeutic effect as a mood stabiliser, lithium exhibits potential anti-ageing effects. This study aimed to examine the relationship between the duration of lithium use, biological ageing and mortality.

**Methods:** The UK Biobank is an observational study of middle-aged and older adults. We tested associations between the duration of lithium use (number of prescriptions, total duration of use, and duration of the first prescription period) and telomere length, frailty, metabolomic age, pulse rate and all-cause mortality.

**Results:** 591 individuals (mean age = 57.49 years; 55% females) had been prescribed lithium. There was no evidence that the number of prescriptions (*β* = −0.022, 95% CI −0.081 to 0.037, *p* = 0.47), the total duration of use (*β* = −0.005, 95% CI −0.023 to 0.013, *p* = 0.57) or the duration of the first prescription period (*β* = −0.018, 95% CI −0.051 to 0.015, *p* = 0.29) correlated with telomere length. There was also no evidence that the duration of lithium use correlated with frailty or metabolomic age. However, a higher prescription count or a longer duration of use was associated with a lower pulse rate. The duration of lithium use did not predict mortality.

**Conclusions:** We observed no evidence of associations between the duration of lithium use and biological ageing markers, including telomere length. Our findings suggest that the potential anti-ageing effects of lithium do not differ by duration of use.

## INTRODUCTION

Lithium is considered the gold standard treatment for patients with bipolar disorder who experience recurrent episodes of (hypo)mania and depression.^1^ Lithium is also used to augment antidepressant medication, for example in patients with difficult-to-treat major depressive disorder.^2^ While the evidence supports lithium’s therapeutic effects,^3^ its mechanisms of action are incompletely understood. Studies indicate that lithium influences neurotransmitter signalling and acts on second messenger systems including the adenyl cyclase and protein kinase A pathways.^4^

Beyond its therapeutic effects as a mood stabiliser, multiple lines of evidence suggest that lithium may impact biological ageing and longevity.^5^ For example, mitochondrial dysfunction is a hallmark of ageing,^6^ and lithium treatment has been shown to enhance mitochondrial metabolism by increasing the activity of electron transport complexes.^7,8^ Preclinical and clinical studies suggest that lithium can reduce oxidative stress, which is linked to several age-related diseases.^9,10^ Notably, studies have demonstrated that lithium extends the lifespan of model organisms, including *Caenorhabditis elegans* (“*C. elegans*”) and Drosophila.^11,12^

A specific focus of research on lithium’s potential anti-ageing properties has been to examine its link with telomere length. Telomeres, which are repetitive nucleoprotein complexes at the ends of chromosomes, progressively shorten with cell division and play an important role in maintaining genomic stability. Telomere attrition is a hallmark of ageing and is linked to cellular senescence.^6^ Telomere dysfunction plays a role in many human diseases.^13^ For example, meta-analyses suggest that individuals with bipolar disorder have shorter telomeres than healthy controls.^14–16^ However, several studies have also found no evidence of a difference in telomere length or observed longer telomeres in patients with bipolar disorder. These inconsistencies may in part result from differences in lithium treatment. Some clinical studies suggest that lithium protects against telomere attrition. For example, studies have shown that bipolar disorder patients treated with lithium had longer telomeres than healthy controls^17^ and that lithium-treated patients had longer telomeres than patients who were not treated with lithium.^17,18^. While these group differences in telomere length could be due to other factors, several studies found that a longer duration of lithium treatment was correlated with longer telomeres.^19^

Building on previous studies in the UK Biobank that have identified predictors and correlates of telomere length,^20–22^ we examined the potential impact of the duration of lithium treatment on telomere length and other markers of biological ageing. We defined three measures of lithium treatment duration using primary care prescription records and assessed multiple markers of biological ageing and longevity, including telomere length, frailty, metabolomic age and all-cause mortality. The overall aim was to elucidate the broader implications of lithium treatment on biological ageing.

## METHODS

### Study population

The UK Biobank is a prospective community-based study that recruited more than 500,000 middle-aged and older adults, aged 40 to 69 years.^23^ Individuals who were registered with the UK National Health Service (NHS) and lived within a 25-mile (∼40 km) radius of one of 22 assessment centres were invited to participate. At the baseline assessment, which took place between 2006 and 2010, participants reported their sociodemographic characteristics, health behaviours and medical history. They also underwent physical examinations and had blood and urine samples taken. Data linkage to primary care health records is available for about 230,000 participants.

### Duration of lithium use

The duration of lithium use was derived from medication prescriptions recorded by general practitioner practices as part of routine patient care. These data included prescriptions recorded between July 1945 and September 2017 and were available for 222,073 participants. Lithium prescriptions were identified using a combination of the British National Formulary (BNF), Read v2 and Dictionary of Medicines and Devices (dm+d) codes, and by searching for relevant string permutations across the prescription medication names. The duration of lithium use was defined in three ways, similar to our previous study on antidepressants.^24^ First, we calculated the total number of lithium prescriptions across all primary care prescription records. Second, we calculated the total number of weeks across all prescription windows, which we defined as periods of less than 90 days between consecutive prescriptions. Third, we calculated the number of weeks of the first prescription window. Prescriptions after the baseline assessment date were excluded from these calculations.

### Telomere length

Leukocyte telomere length was measured using a quantitative polymerase chain reaction (PCR) assay that expresses telomere length as the ratio of the telomere repeat copy number (T) relative to a single-copy gene (S) that encodes haemoglobin subunit beta.^25^ The T/S ratio is proportional to an individual’s average telomere length.^26^ Measurements were adjusted for operational and technical parameters (PCR machine, staff member, enzyme batch, primer batch, temperature, humidity, primer batch × PCR machine, primer batch × staff member, A260/A280 ratio of the DNA sample and A260/A280 ratio squared), log_e_ transformed and Z-standardised.

### Frailty index

A frailty index was derived from health deficits that met the following criteria: indicators of poor health; more prevalent in older individuals; neither rare nor universal; covering multiple areas of functioning; available for ≥ 80% of participants.^27^ The index included 49 variables ascertained via touch-screen questionnaires and nurse-led interviews, including cardiometabolic, cranial, immunological, musculoskeletal, respiratory, and sensory traits, well-being, infirmity, cancer, and pain. Categorical variables were dichotomised (no deficit = 0; deficit = 1), and ordinal variables were mapped onto a score between zero and one. The sum of deficits was divided by the total number of possible deficits, resulting in frailty index scores between zero and one, with higher scores indicating greater levels of frailty.^28^ Participants with missing data for ≥ 10/49 variables were excluded.^27^

### Metabolomic ageing clock

Nuclear magnetic resonance (NMR) spectroscopy-derived metabolomic biomarkers were quantified in non-fasting plasma samples using the Nightingale Health platform, which ascertains 168 circulating metabolites using a high-throughput standardised protocol.^29^ In 101,359 participants, we developed a metabolomic ageing clock using a Cubist rule-based regression model. This algorithm is an ensemble technique that derives rules from decision trees and fits linear regression models in the subsets of data defined by these rules. The model incorporates boosting techniques and may adjust predictions based on *k*-nearest neighbors.^30,31^ Model performance was assessed using nested cross-validation with 10 outer loops and 5 inner folds for hyperparameter tuning. The cross-validation mean absolute error (MAE) was 5.42 years. Metabolomic age (MileAge) delta represents the difference between predicted and chronological age, with positive and negative values indicating accelerated and decelerated biological ageing, respectively.

### Pulse rate

Resting pulse rate in beats per minute was recorded using an Omron 705 IT digital blood pressure monitor device or, exceptionally, a manual sphygmomanometer. We calculated the average of the two available readings to reduce potential measurement error.

### All-cause mortality

The date of death was obtained through linkage with national death registries: NHS Digital (England and Wales) and the NHS Central Register (Scotland). The censoring date was 30 November 2022. The most recent death was recorded on 19 December 2022, although the data were incomplete for December 2022.

### Covariates

Covariates for the cross-sectional and prospective analyses included chronological age, sex, body mass index (kg/m²), morbidity count (none/one, two, three, four or five+), gross annual household income (<£18,000, £18,000–£30,999, £31,000–£51,999 or £52,000–£100,000 / >£100,000), highest educational/professional qualification (none, O levels/GCSEs/CSEs, A levels/NVQ/HND/HNC or degree) and cohabitation status (single or with spouse/partner).

### Statistical analyses

The data processing and analyses were performed in R (version 4.2). Sample characteristics were summarised using means and standard deviations or counts and percentages. Associations between the duration of lithium use (number of prescriptions, total duration of use, and duration of the first prescription period) and telomere length were estimated using ordinary least squares regression. We further tested associations between the duration of lithium use and two other indicators of biological age, the frailty index and the metabolomic ageing clock, and pulse rate. Hazard ratios (HRs) and 95% confidence intervals were estimated using Cox proportional hazards models to examine associations between the duration of lithium use and all-cause mortality. We examined both quintiles of the distribution, with the lowest quintile as the reference group, and spline functions of the distribution with the median as reference value. Age in years was used as the underlying time axis, with age 40 as the start of follow-up. The cross-sectional and prospective analyses were adjusted for chronological age and sex (Model 1) and chronological age, sex, body mass index, morbidity count, household income, highest qualification, and cohabitation status (Model 2).

### Sensitivity analyses

We conducted the following sensitivity analyses of the duration of lithium use and biological ageing markers: we restricted the sample to individuals with a total duration of lithium use of (a) at least 1 year (52 weeks) or (b) at least 4.5 years (234 weeks); we additionally adjusted for current antidepressant medication use (yes/no) at the time of the baseline assessment.

## RESULTS

### Sample characteristics

Of the 502,476 participants in the UK Biobank, 222,073 individuals (about 44.2% of the sample) had prescription data available through their linked primary care records. Of the 773 participants with a lithium prescription in their primary care record, *n* = 182 were excluded because their first prescription followed the baseline assessment. The duration of lithium use was estimated for 591 participants (Figure S1). Sample characteristics are shown in Table 1. Based on our previously reported criteria,^32,33^ 71.74% of participants had a lifetime history of mood disorders (*n* = 248 had bipolar disorder and *n* = 176 had depression).

**Table 1.** Sample characteristics.

| | Lithium duration<br>($N=591$) | Primary care<br>($N=221,482$) | No primary care<sup>1</sup><br>($N=280,403$) |
| --- | --- | --- | --- |
| **Age; mean (SD)** | 57.49 (7.51) | 56.58 (8.05) | 56.48 (8.13) |
| **Sex** |  |  |  |
| Female | 327 (55.3%) | 121618 (54.9%) | 151420 (54.0%) |
| Male | 264 (44.7%) | 99864 (45.1%) | 128982 (46.0%) |
| **Ethnicity** |  |  |  |
| White | 572 (96.8%) | 210119 (94.9%) | 261977 (93.4%) |
| Non-White | 14 (2.4%) | 10322 (4.7%) | 16695 (6.0%) |
| Missing<sup>2</sup> | 5 (0.8%) | 1041 (0.5%) | 1731 (0.6%) |
| **Highest qualification** |  |  |  |
| None | 119 (20.1%) | 39882 (18.0%) | 45265 (16.1%) |
| O levels/GCSEs/CSEs | 125 (21.2%) | 58292 (26.3%) | 73662 (26.3%) |
| A levels/NVQ/HND/HNC<sup>3</sup> | 118 (20.0%) | 50521 (22.8%) | 63207 (22.5%) |
| Degree | 222 (37.6%) | 69916 (31.6%) | 91014 (32.5%) |
| Missing<sup>2</sup> | 7 (1.2%) | 2871 (1.3%) | 7255 (2.6%) |
| **Household income<sup>4</sup>** |  |  |  |
| Very low | 198 (33.5%) | 45278 (20.4%) | 51717 (18.4%) |
| Low | 132 (22.3%) | 49247 (22.2%) | 58794 (21.0%) |
| Medium | 95 (16.1%) | 49037 (22.1%) | 61637 (22.0%) |
| High / Very high | 72 (12.2%) | 45305 (20.5%) | 63809 (22.8%) |
| Missing<sup>2</sup> | 94 (15.9%) | 32615 (14.7%) | 44446 (15.9%) |
| **Cohabitation** |  |  |  |
| With partner | 359 (60.7%) | 160735 (72.6%) | 201961 (72.0%) |
| Single | 55 (9.3%) | 17885 (8.1%) | 24172 (8.6%) |
| Missing<sup>2</sup> | 177 (29.9%) | 42862 (19.4%) | 54270 (19.4%) |
| **Smoking status** |  |  |  |
| Never | 293 (49.6%) | 121023 (54.6%) | 152188 (54.3%) |
| Former | 186 (31.5%) | 76153 (34.4%) | 96711 (34.5%) |
| Current | 106 (17.9%) | 23143 (10.4%) | 29724 (10.6%) |
| Missing<sup>2</sup> | 6 (1.0%) | 1163 (0.5%) | 1780 (0.6%) |
| **Morbidity count** |  |  |  |
| None / One | 119 (20.1%) | 108494 (49.0%) | 140930 (50.3%) |
| Two | 150 (25.4%) | 44962 (20.3%) | 55724 (19.9%) |
| Three | 116 (19.6%) | 29465 (13.3%) | 36037 (12.9%) |
| Four | 77 (13.0%) | 17297 (7.8%) | 20903 (7.5%) |
| Five+ | 126 (21.3%) | 20903 (9.4%) | 26310 (9.4%) |
| Missing<sup>2</sup> | 3 (0.5%) | 361 (0.2%) | 499 (0.2%) |
| **Baseline lithium Rx<sup>5</sup>** |  |  |  |
| No | 296 (50.1%) | 204499 (92.3%) | 252446 (90.0%) |
| Yes | 261 (44.2%) | 83 (0.0%) | 417 (0.1%) |
| Missing<sup>2</sup> | 34 (5.8%) | 16900 (7.6%) | 27540 (9.8%) |
| **Baseline cholesterol Rx<sup>5</sup>** |  |  |  |
| No | 425 (71.9%) | 171354 (77.4%) | 217473 (77.6%) |
| Yes | 161 (27.2%) | 48000 (21.7%) | 57335 (20.4%) |
| Missing<sup>2</sup> | 5 (0.8%) | 2128 (1.0%) | 5595 (2.0%) |
| **Fasting time** |  |  |  |
| Less than 8 hours | 563 (95.3%) | 211768 (95.6%) | 267565 (95.4%) |
| At least 8 hours | 24 (4.1%) | 9179 (4.1%) | 12143 (4.3%) |
| Missing<sup>2</sup> | 4 (0.7%) | 535 (0.2%) | 695 (0.2%) |
| **Body mass index; mean (SD)<sup>6</sup>** | **28.70 (5.34)** | **27.54 (4.83)** | **27.34 (4.78)** |

*Note:* Numbers shown are counts and percentages unless indicated otherwise. SD = standard deviation; GCSEs = general certificate of secondary education; CSE = certificate of secondary education; NVQ = national vocational qualification; HND = higher national diploma; HNC = higher national certificate; Rx = prescription. <sup>1</sup> Information on age and sex was missing for one participant in the sample without linked primary care data. <sup>2</sup> Missing data also includes “do not know”, “prefer not to answer” or “not applicable”. <sup>3</sup> Also includes 'other professional qualifications'. <sup>4</sup> Annual household income groups: very low (<£18,000), low (£18,000–£30,999), middle (£31,000–£51,999), high (£52,000–£100,000) and very high (>£100,000). <sup>5</sup> Prescriptions refer to *current* medication use at the time of the baseline assessment. <sup>6</sup> Missing data for body mass index: $n = 10$ , $n = 1344$ and $n = 1751$ .

### Duration of lithium use and telomere length

In the model adjusted for chronological age and sex, there was no evidence that the number of lithium prescriptions (*N* = 569, *β* = −0.022, 95% CI −0.081 to 0.037, *p* = 0.47), the total duration of use (*N* = 569, *β* = −0.005, 95% CI −0.023 to 0.013, *p* = 0.57) or the duration of the first prescription period (*N* = 569, *β* = −0.018, 95% CI −0.051 to 0.015, *p* = 0.29) were associated with telomere length. Additional adjustment for body mass index, morbidity count, household income, highest qualification and cohabitation status had no discernible impact on these results (Table 1). Individuals with a higher prescription count or longer duration of lithium use were more likely to use lithium at the time of the baseline assessment (Figure 1).

**Figure 1.**
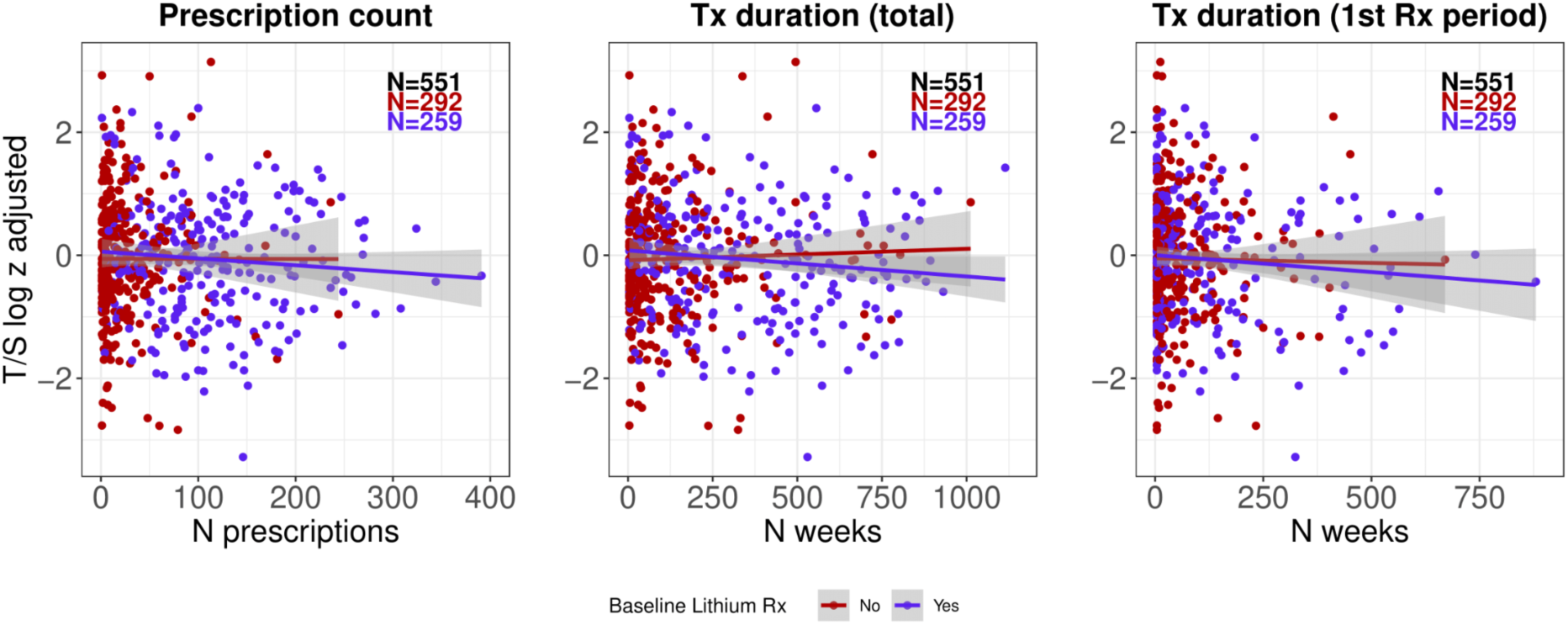
Scatter plots of telomere length (T/S ratio log_e_ transformed and Z standardised) by lithium prescription count, total duration of use, and duration of the first prescription period. The lines were estimated using ordinary least squares regression, and the shaded areas correspond to 95% confidence intervals. Lithium use (yes/no) at the time of the baseline assessment when telomere length was measured is shown in different colours. Individuals without baseline data on current lithium use are not shown. Tx = treatment; Rx = prescription.

### Duration of lithium use, frailty and metabolomic age

There was no evidence that the duration of lithium use was associated with the frailty index (*N* = 585, *β* = −0.012, 95% CI −0.030 to 0.006, *p* = 0.21) or the metabolomic ageing clock (*N* = 120, *β* = 0.030, 95% CI −0.133 to 0.194, *p* = 0.72) (Figure 2). This finding was consistent across all measures of the duration of lithium use and adjustment for additional covariates (Table 1).

**Figure 2.**
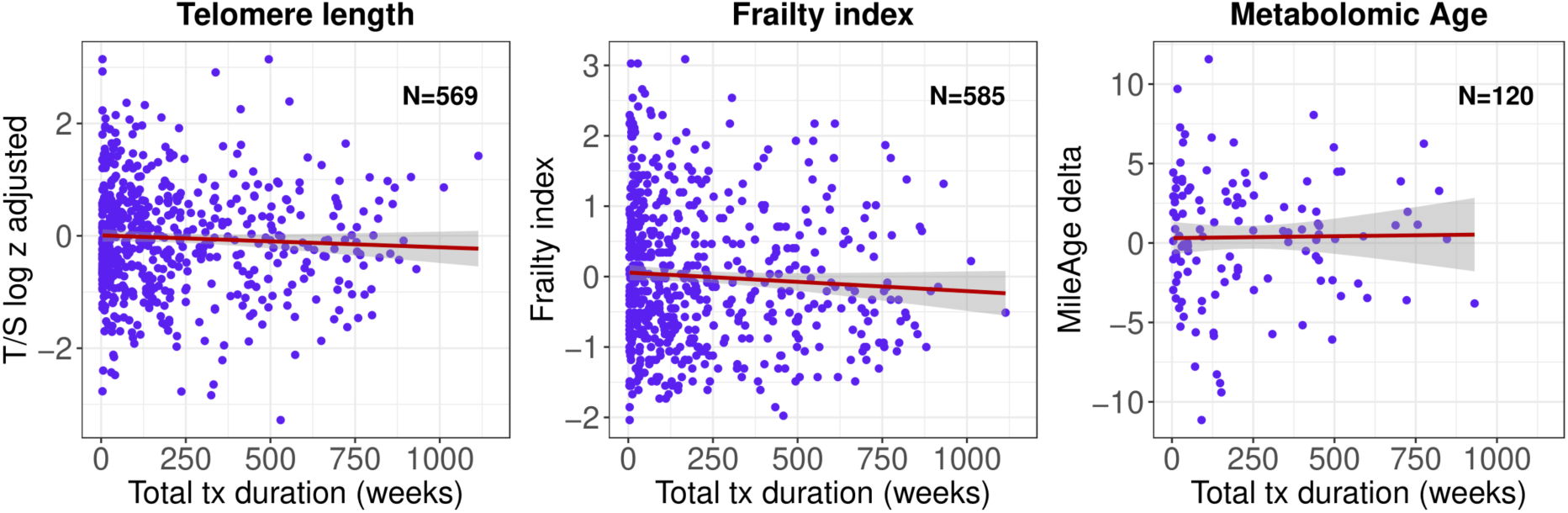
Scatter plots of telomere length (T/S ratio log_e_ transformed and Z standardised), the frailty index, and the metabolomic ageing clock delta by total duration of lithium use. The lines were estimated using ordinary least squares regression, and the shaded areas correspond to 95% confidence intervals. Tx = treatment.

**Table 2.** Associations between the duration of lithium use and biological ageing markers Model 1 (adj. for age and sex) Model 2 (full adjustment)

|  | Model 1 (adj. for age and sex) |  |  |  | Model 2 (full adjustment) |  |  |  |
| --- | --- | --- | --- | --- | --- | --- | --- | --- |
|  | *n* | Beta | 95% CI | *p* | *n* | Beta | 95% CI | *p* |
| **Telomere length** |  |  |  |  |  |  |  |  |
| Rx count | 569 | -0.022 | -0.081 0.037 | 0.47 | 567 | -0.017 | -0.076 0.043 | 0.58 |
| Total duration | 569 | -0.005 | -0.023 0.013 | 0.57 | 567 | -0.004 | -0.022 0.015 | 0.71 |
| Duration first Rx | 569 | -0.018 | -0.051 0.015 | 0.29 | 567 | -0.014 | -0.048 0.019 | 0.40 |
| **Frailty index** |  |  |  |  |  |  |  |  |
| Rx count | 585 | -0.045 | -0.104 0.014 | 0.13 | 580 | -0.043 | -0.091 0.004 | 0.08 |
| Total duration | 585 | -0.012 | -0.030 0.006 | 0.21 | 580 | -0.013 | -0.028 0.001 | 0.07 |
| Duration first Rx | 585 | -0.027 | -0.060 0.007 | 0.12 | 580 | -0.021 | -0.048 0.007 | 0.14 |
| **Metabolomic age** |  |  |  |  |  |  |  |  |
| Rx count | 120 | -0.036 | -0.590 0.518 | 0.90 | 120 | -0.022 | -0.583 0.540 | 0.94 |
| Total duration | 120 | 0.030 | -0.133 0.194 | 0.72 | 120 | -0.009 | -0.180 0.161 | 0.91 |
| Duration first Rx | 120 | 0.248 | -0.079 0.576 | 0.14 | 120 | 0.208 | -0.128 0.543 | 0.22 |
| **Pulse rate** |  |  |  |  |  |  |  |  |
| Rx count | 586 | -0.061 | -0.119 -0.002 | 0.04 | 583 | -0.058 | -0.116 0.0001 | 0.05 |
| Total duration | 586 | -0.026 | -0.044 -0.008 | 0.005 | 583 | -0.024 | -0.042 -0.006 | 0.01 |
| Duration first Rx | 586 | -0.021 | -0.055 0.012 | 0.21 | 583 | -0.022 | -0.056 0.011 | 0.18 |

*Note:* The measurement units have been rescaled. The number of prescriptions were analysed in units of 50 prescriptions; the total duration of use and the duration of the first prescription period were expressed in years. Model 1 was adjusted for age and sex; Model 2 was adjusted for age, sex, body mass index, morbidity count, household income, highest qualification and cohabitation status. Rx = prescription; CI = confidence interval.

**Table 3.** Associations between the duration of lithium use and all-cause mortality Model 1 (adj. for age and sex) Model 2 (full adjustment)

|  |  | Model 1 (adj. for age and sex) |  |  |  |  |  | Model 2 (full adjustment) |  |  |  |  |  |
| --- | --- | --- | --- | --- | --- | --- | --- | --- | --- | --- | --- | --- | --- |
|  |  | N<sub>total</sub> | N<sub>died</sub> | HR | 95% CI |  | *p* | N<sub>total</sub> | N<sub>died</sub> | HR | 95% CI |  | *p* |
| Rx count |  |  |  |  |  |  |  |  |  |  |  |  |  |
| 1st | ≤ 7 | 119 | 19 | Reference |  |  |  | 118 | 19 | Reference |  |  |  |
| 2nd | 8-23 | 118 | 16 | 0.68 | 0.34 | 1.36 | 0.28 | 116 | 15 | 0.53 | 0.26 | 1.09 | 0.08 |
| 3rd | 24-60 | 118 | 19 | 0.88 | 0.46 | 1.69 | 0.70 | 117 | 19 | 0.74 | 0.37 | 1.48 | 0.40 |
| 4th | 61-114 | 118 | 18 | 0.71 | 0.37 | 1.38 | 0.32 | 118 | 18 | 0.68 | 0.34 | 1.35 | 0.27 |
| 5th | ≥ 115 | 118 | 25 | 1.05 | 0.57 | 1.93 | 0.88 | 116 | 25 | 0.91 | 0.48 | 1.74 | 0.79 |
| Total duration |  |  |  |  |  |  |  |  |  |  |  |  |  |
| 1st | ≤ 22 | 119 | 19 | Reference |  |  |  | 118 | 19 | Reference |  |  |  |
| 2nd | 23-83 | 118 | 14 | 0.77 | 0.38 | 1.54 | 0.45 | 117 | 13 | 0.61 | 0.29 | 1.26 | 0.18 |
| 3rd | 84-187 | 118 | 19 | 0.83 | 0.43 | 1.62 | 0.59 | 115 | 19 | 0.74 | 0.37 | 1.49 | 0.40 |
| 4th | 188-426 | 118 | 21 | 1.15 | 0.61 | 2.17 | 0.66 | 118 | 21 | 0.95 | 0.50 | 1.83 | 0.88 |
| 5th | ≥ 427 | 118 | 24 | 1.01 | 0.55 | 1.86 | 0.98 | 117 | 24 | 0.91 | 0.48 | 1.73 | 0.78 |
| Duration first Rx |  |  |  |  |  |  |  |  |  |  |  |  |  |
| 1st | ≤ 4 | 119 | 21 | Reference |  |  |  | 117 | 20 | Reference |  |  |  |
| 2nd | 5-16 | 118 | 17 | 1.04 | 0.54 | 2.00 | 0.90 | 118 | 17 | 0.96 | 0.49 | 1.92 | 0.92 |
| 3rd | 17-49 | 118 | 13 | 0.74 | 0.36 | 1.53 | 0.42 | 117 | 13 | 0.71 | 0.34 | 1.50 | 0.37 |
| 4th | 50-130 | 118 | 19 | 0.95 | 0.51 | 1.79 | 0.88 | 117 | 19 | 1.05 | 0.54 | 2.03 | 0.90 |
| 5th | ≥ 131 | 118 | 27 | 1.48 | 0.82 | 2.66 | 0.19 | 116 | 27 | 1.46 | 0.80 | 2.68 | 0.22 |

*Note:* Hazard ratios and 95% confidence intervals from Cox proportional hazards models for all-cause mortality. Age (in years) was used as the underlying time axis. Model 1 was adjusted for age and sex; Model 2 was adjusted for age, sex, body mass index, morbidity count, household income, highest qualification and cohabitation status. Rx = prescription; HR = hazard ratio; CI = confidence interval.

### Duration of lithium use and pulse rate

To ensure that the lack of statistically significant associations was not due to how we defined the duration of lithium use, we further tested associations between the duration of lithium use and pulse rate (Figure S2). The number of lithium prescriptions (*N* = 586, *β* = −0.061, 95% CI −0.119 to −0.002, *p* = 0.04) and the total duration of lithium use (*N* = 586, *β* = −0.026, 95% CI - 0.044 to −0.008, *p* = 0.005) were both associated with a lower pulse rate (Table 1).

### Duration of lithium use and all-cause mortality

The median duration of follow-up of censored individuals was 13.71 years (IQR = 1.35 years), with more than 7686 person-years of follow-up. During this period, 94 deaths were observed. There was no evidence that the duration of lithium use predicted all-cause mortality. Comparing individuals in the second to fifth quintile of the distribution of the duration of lithium use to individuals in the first quintile resulted in no statistically significant differences in all-cause mortality (lowest *p* = 0.08) (Figure 3).

**Figure 3.**
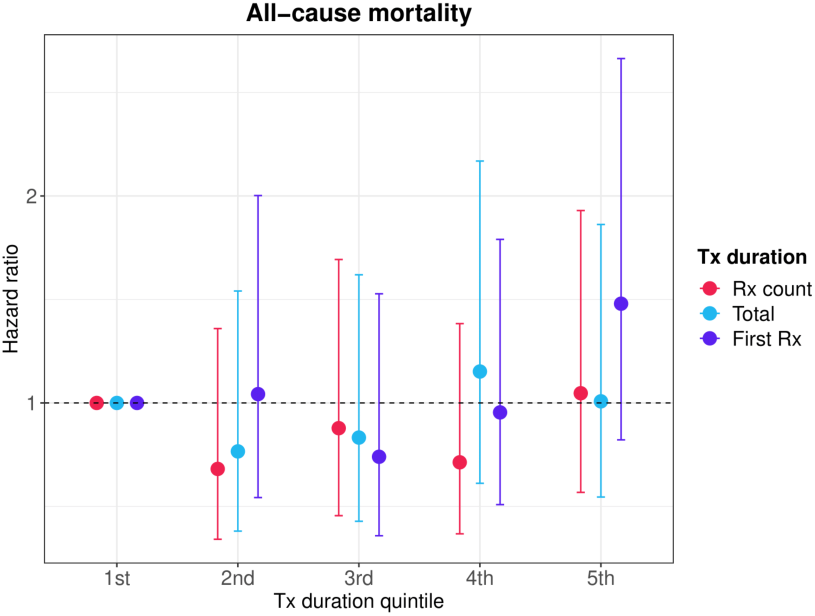
Hazard ratios (HR) and 95% confidence intervals from Cox proportional hazards models for all-cause mortality. Age (in years) was used as the underlying time axis. The model was adjusted for age and sex. Reference group: individuals in the lowest quintile of the distribution of the duration of lithium use (prescription count, total duration and duration of the first prescription period). Tx = treatment; Rx = prescription.

Modelling the mortality hazard as a spline function of the duration of lithium use, instead of by quintiles of the distribution, provided little evidence of a statistically significant association between the duration of lithium use and all-cause mortality (Figure 4 and Figure S3). There was, however, some evidence that a prescription count greater than about 300 was associated with a higher mortality risk.

**Figure 4.**
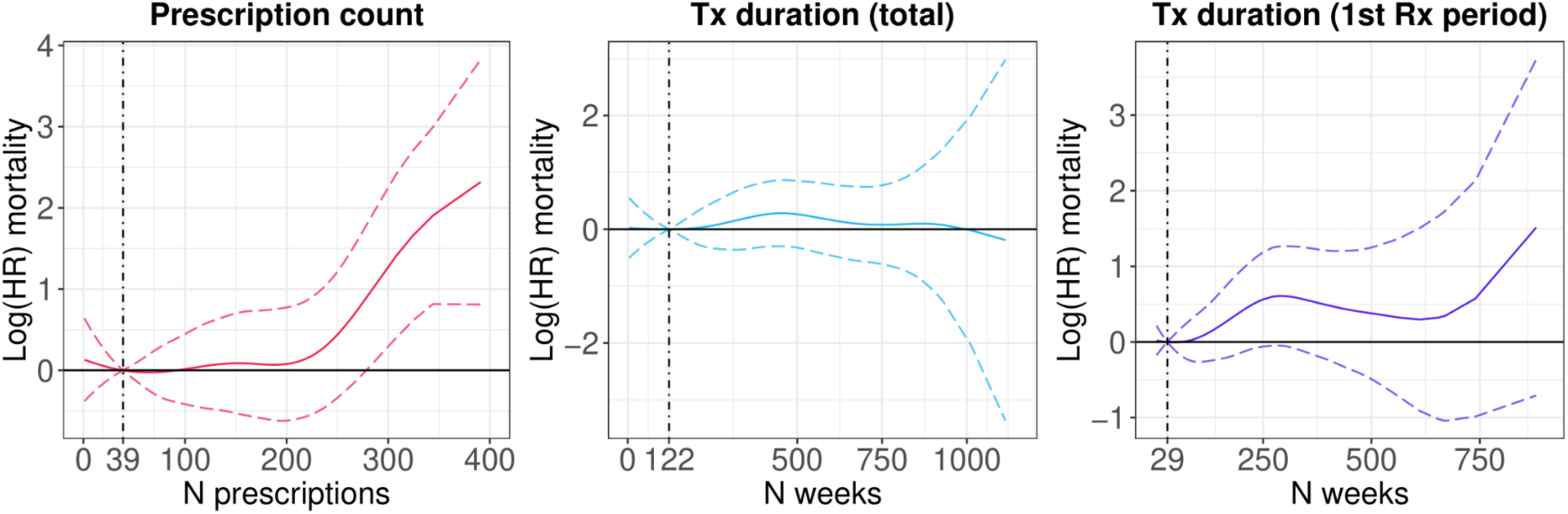
Log hazard ratios and 95% confidence intervals from Cox proportional hazards models for all-cause mortality. Age (in years) was used as the underlying time axis. The model was adjusted for age and sex. Vertical lines indicate the median of the distribution. Tx = treatment; Rx = prescription.

### Sensitivity analyses

Excluding participants with a total duration of lithium use of less than 1 year or less than 4.5 years provided little evidence that the duration of lithium use was associated with biological ageing (Table S1 and Table S2). There was some evidence that a longer duration of the first lithium prescription period was associated with a higher MileAge delta, although this finding did not survive multiple testing correction in the fully adjusted model. The duration of lithium use was also not associated with pulse rate in these analyses.

The relationship between telomere length and the duration of lithium use stratified by current antidepressant use at the time of the baseline assessment is shown in Figure S4. Additional adjustment for current antidepressant use had no discernible impact on the analyses of the relationship between the duration of lithium use and biological ageing (Table S3). However, the association between a higher prescription count and a longer total duration of lithium use and pulse rate was stronger in these analyses.

## DISCUSSION

We examined the relationship between the duration of lithium use and multiple markers of biological ageing, including telomere length, frailty, metabolomic age, and all-cause mortality. Both cross-sectional and prospective analyses provided little evidence of an association between the duration of lithium use and biological ageing or mortality.

Our findings are inconsistent with most prior studies of the duration of lithium use and telomere length, several of which identified a positive association between lithium and telomere length. Martinsson et al. found that lithium-treated bipolar disorder patients had on average 35% longer telomeres than healthy controls.^17^ In the same study, the duration of lithium treatment was positively correlated with telomere length, particularly in patients who had been treated for more than 2.5 years. Pisanu et al.^34^ and Coutts et al.^19^ corroborated these observations, identifying statistically significant associations between chronic lithium treatment and telomere length in patients with bipolar disorder. Squassina et al. also reported a positive correlation between the duration of lithium use and telomere length in 150 bipolar disorder patients who had been treated for more than two years.^35^ A mechanistic *in vitro* study provided further evidence supporting lithium’s putative effect on telomeres by demonstrating that lithium administration to lymphoblastoid cell lines from bipolar disorder patients increased telomere length.^18^ Nevertheless, there is some precedent for our findings. Ferensztajn-Rochowiak et al. reported that in a sample of 41 patients with bipolar disorder, the duration of lithium treatment did not correlate with telomere length in peripheral blood leukocytes.^36^ Another *in vitro* study found that administration of lithium to lymphoblastoid cell lines from bipolar disorder patients had no effect on telomere length,^37^ though it is worth noting that these patients had previously been treated with lithium.

There are several potential explanations for the observed differences between our findings and those of most prior studies. For example, most clinical studies determined the duration of lithium treatment retrospectively during the study visit, while we derived the duration of lithium use in our study from prescription records. Furthermore, there is some evidence that clinical factors such as bipolar disorder diagnosis, number of episodes, chronicity and severity of illness impact telomere length. The participants in our study likely represent a healthier subset of the population than the patients included in prior clinical studies. Moreover, genetic factors can moderate the effect of lithium on telomere length.^19^ Although, we adjusted for several confounders in our analyses, we also found little evidence of any statistical associations in the model adjusted only for chronological age and sex. The sample size of our study (*n* = 569) is one of the largest in the field, and we used three definitions of the duration of lithium use to increase the robustness of our findings. In addition, we conducted a sensitivity analysis of pulse rate, which we expected to differ with longer duration of lithium use.^38^ The observed correlation between the duration of lithium use and a lower pulse rate supports the validity of our measures of the duration of lithium use.

There has been little prior research on the relationship between lithium use and the other biological ageing markers explored in this study. Okazaki et al. found that bipolar disorder patients who were taking mood stabilisers had a younger epigenetic age, according to Horvath’s epigenetic ageing clock, than patients who were not taking mood stabilisers.^39^ However, an *in vitro* study of lymphoblastoid cell lines did not find that administration of lithium impacted the Horvath clock.^18^ We found no evidence of an association between the duration of lithium use and metabolomic age in our study. However, it is worth noting that epigenetic and metabolomic age are not strongly correlated.^40^ We also explored the relationship between the duration of lithium use and frailty, which is a measure of age-related health deficit accumulation. While some evidence suggests that frailty and telomere length are not strongly linked,^41^ frailty is a key marker of biological ageing and physical ill-health.^42,43^ Our study provided no evidence of a statistically significant association between the duration of lithium use and frailty. Taken together, these findings suggest that the lack of association between the duration of lithium use and biological ageing – at least in our sample – was not specific to telomere length.

The *duration* of lithium use was not linked to all-cause mortality in our study. Multiple observational studies have previously shown that lithium use is associated with a lower mortality risk. For example, a cohort study of 826 patients in Finland found that lithium treatment, compared to no lithium use, was associated with a lower all-cause mortality risk.^44^ Another cohort study in Taiwan found that lithium was associated with the lowest all-cause mortality risk in bipolar disorder patients, relative to other mood stabiliser treatments.^45^ A recent study in the UK Biobank also found that lithium treatment was associated with a lower all-cause mortality risk than treatment with antipsychotics.^46^ However, it is important to note that we examined the duration of lithium use, not whether patients were treated versus not treated with lithium, which prior studies examined. As such, our findings suggest that there may not be a link between longer duration of lithium use and all-cause mortality in a community-based, real-world sample. These findings do not imply that lithium use (yes/no) is not linked to all-cause mortality, and they do not invalidate findings from clinical trials where lithium adherence is more controlled and all-cause mortality.

### Limitations

We acknowledge certain limitations to our study. The UK Biobank primarily recruited individuals between the ages of 40 to 69 years, limiting the conclusions drawn from our findings to middle-aged and older adults. There is evidence of a healthy volunteer participation bias in the UK Biobank,^47,48^ which could mean that the participants in our study were healthier than the patients included in prior clinical studies. The observational nature of our study precludes any causal inferences, and residual confounding factors may exist despite our efforts to account for relevant covariates. The duration of lithium use was derived from primary care prescription records. Prescriptions may not perfectly reflect medication use and there is some evidence that, despite the need for regular blood tests, adherence to lithium is poor in clinical practice;^49^ furthermore, prescription dates are a proxy of date of use. Nonetheless, we derived and tested three different definitions of the duration of lithium use to be as robust as possible within these constraints. Telomere length was measured using leukocyte DNA, and findings might differ from telomere length measured in other tissues. However, prior evidence suggests that leukocyte telomere length correlates well with telomere length in other tissues.^50^ Finally, we could only examine *average* telomere length and were unable to evaluate whether the duration of lithium use was associated with variability in telomere length, which may be more important for telomere dysfunction and biological ageing.^26^

## Conclusion

Our findings, in a larger sample, challenge prior research on the relationship between the duration of lithium use and telomere length. Given that we also did not observe statistically significant associations with other biological ageing markers such as frailty and metabolomic age, nor with all-cause mortality, our findings suggest that the potential anti-ageing effects of lithium do not differ by duration of use. Our findings encourage further scientific inquiry into the complex interplay between lithium medication, biological ageing and lifespan.

## ETHICS

Ethical approval for the UK Biobank study has been granted by the National Information Governance Board for Health and Social Care and the NHS North West Multicentre Research Ethics Committee (11/NW/0382). No project-specific ethical approval is needed.

## AUTHORSHIP CONTRIBUTIONS

JM conceived the idea of the study, acquired the data, carried out the statistical analysis, interpreted the findings and co-wrote the manuscript. WLEW identified the lithium medication codes, pre-processed the data to define duration of lithium use, interpreted the findings and co-wrote the manuscript. TRP, AHY, GSD and CML interpreted the findings and reviewed the manuscript. All authors read and approved the final manuscript.

## Data Availability

UK Biobank data used are available to all bona fide researchers for health-related research that is in the public interest, subject to an application process and approval criteria. Study materials are publicly available online at http://www.ukbiobank.ac.uk.

## ACKNOWLEDGMENTS

This study was funded by the National Institute for Health and Care Research (NIHR) Maudsley Biomedical Research Centre at South London and Maudsley NHS Foundation Trust and King’s College London. The views expressed are those of the authors and not necessarily those of the NHS, the NIHR or the Department of Health and Social Care. WLEW is supported by the National University of Singapore President’s Graduate Fellowship. TRP is supported by the MRC (UKRI) as part of a New Investigator Research Grant [MR/W028018/1]. GSD is supported by the Ministry of Education, Singapore, under its Academic Research Fund Tier 3 Award (MOE2017-T3-1-002). This research has been conducted using data from UK Biobank, a major biomedical database. Data access permission has been granted under UK Biobank application 45514. All analyses were supported by: King’s College London. (2023). King’s Computational Research, Engineering and Technology Environment (CREATE). Retrieved May 24, 2023, from https://doi.org/10.18742/rnvf-m076

## DATA SHARING STATEMENT

UK Biobank data used are available to all *bona fide* researchers for health-related research that is in the public interest, subject to an application process and approval criteria. Study materials are publicly available online at http://www.ukbiobank.ac.uk.

