## Supplementary material for "The duration of lithium use and biological ageing: telomere length, frailty, metabolomic age and all-cause mortality": Figure S1; Figure S2; Figure S3; Figure S4; Table S1; Table S2; Table S3

This material accompanies the article

### Distribution of lithium use duration

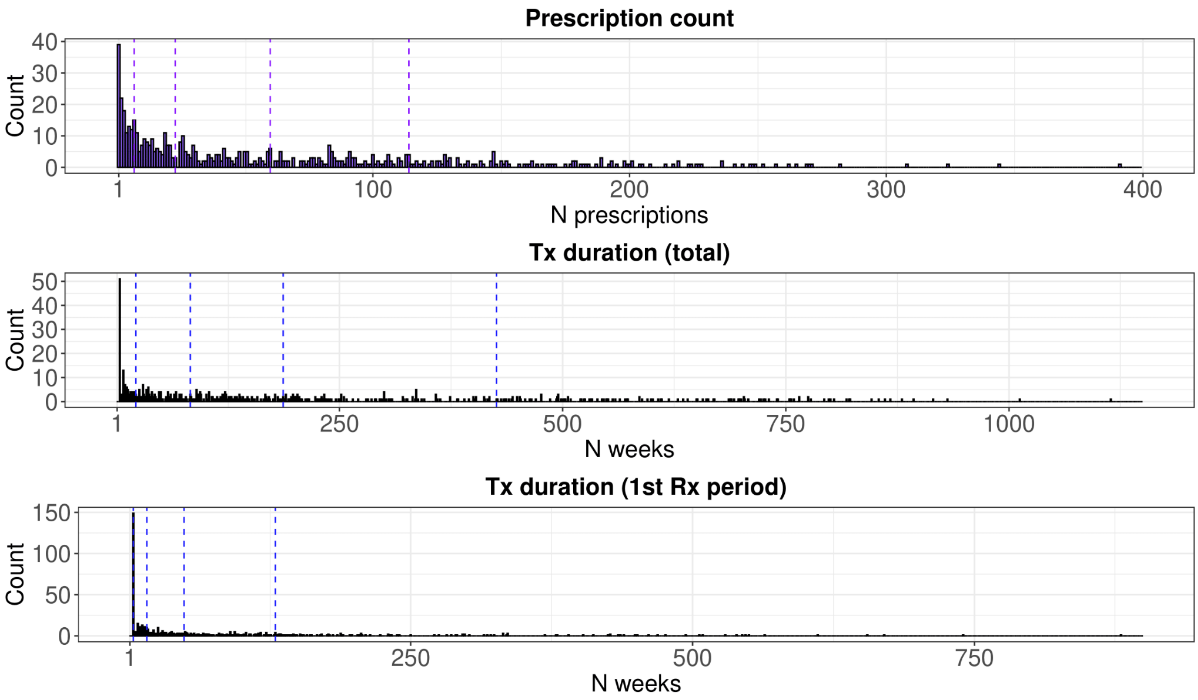

**Figure S1.** Histograms of lithium prescription count, total duration of use and duration of the first prescription period. Vertical lines indicate quintiles of the distribution. Tx = treatment; Rx = prescription.

Lithium use duration and pulse rate

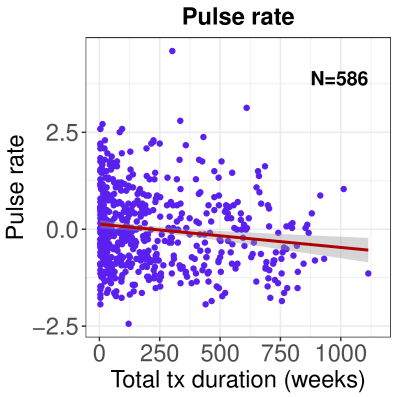

**Figure S2.** Scatter plot of pulse rate by total duration of lithium use. The lines were estimated using ordinary least squares regression and the shaded areas correspond to 95% confidence intervals. Tx = treatment.

Lithium use duration and mortality

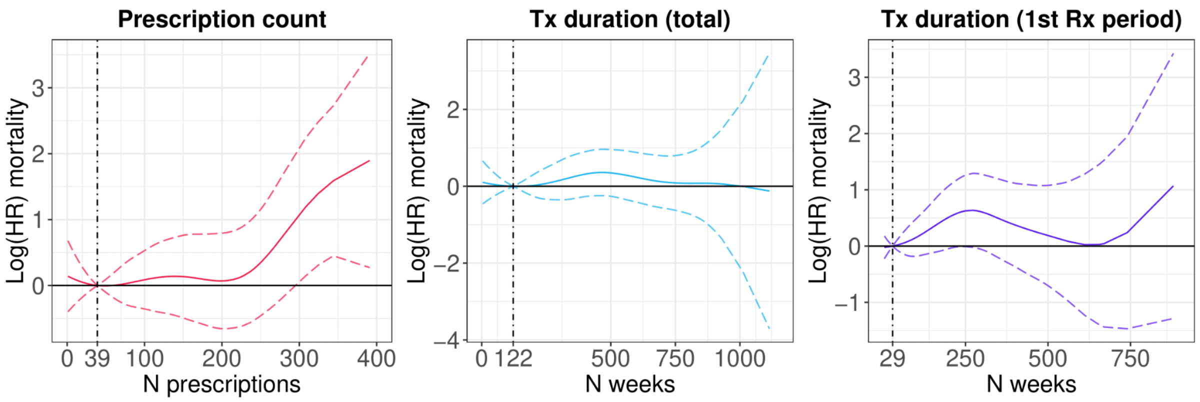

**Figure S3.** Log hazard ratios and 95% confidence intervals from Cox proportional hazards models for all-cause mortality. Age (in years) was used as the underlying time axis. The model was adjusted for age, sex, body mass index, morbidity count, household income, highest qualification, and cohabitation status. Vertical lines indicate the median of the distribution. Tx = treatment; Rx = prescription.

Sensitivity analysis: minimum duration 1 year

**Table S1.** Associations between the duration of lithium use and biological ageing markers, excluding individuals with a total duration of lithium use < 1 year

| | Model 1 (adj. for age and sex) | | | | | Model 2 (full adjustment) | | | | |
|---|---|---|---|---|---|---|---|---|---|---|
| | *n* | Beta | 95% CI | | *p* | *n* | Beta | 95% CI | | *p* |
| **Telomere length** | | | | | | | | | | |
| Rx count | 380 | -0.028 | -0.098 | 0.041 | 0.43 | 378 | -0.022 | -0.093 | 0.050 | 0.55 |
| Total duration | 380 | -0.005 | -0.027 | 0.017 | 0.64 | 378 | -0.003 | -0.026 | 0.019 | 0.76 |
| Duration first Rx | 380 | -0.019 | -0.054 | 0.017 | 0.30 | 378 | -0.015 | -0.051 | 0.022 | 0.43 |
| **Frailty index** | | | | | | | | | | |
| Rx count | 389 | -0.016 | -0.085 | 0.052 | 0.64 | 385 | -0.016 | -0.072 | 0.041 | 0.59 |
| Total duration | 389 | -0.001 | -0.022 | 0.021 | 0.94 | 385 | -0.003 | -0.021 | 0.015 | 0.73 |
| Duration first Rx | 389 | -0.018 | -0.053 | 0.017 | 0.32 | 385 | -0.009 | -0.038 | 0.020 | 0.54 |
| **Metabolomic age** | | | | | | | | | | |
| Rx count | 81 | 0.333 | -0.421 | 1.086 | 0.38 | 81 | 0.283 | -0.552 | 1.117 | 0.50 |
| Total duration | 81 | 0.152 | -0.073 | 0.376 | 0.18 | 81 | 0.063 | -0.196 | 0.322 | 0.63 |
| Duration first Rx | 81 | 0.389 | 0.013 | 0.765 | 0.04 | 81 | 0.335 | -0.078 | 0.748 | 0.11 |
| **Pulse rate** | | | | | | | | | | |
| Rx count | 391 | -0.038 | -0.108 | 0.033 | 0.29 | 388 | -0.036 | -0.108 | 0.035 | 0.32 |
| Total duration | 391 | -0.019 | -0.042 | 0.003 | 0.09 | 388 | -0.017 | -0.040 | 0.005 | 0.14 |
| Duration first Rx | 391 | -0.007 | -0.043 | 0.029 | 0.70 | 388 | -0.014 | -0.050 | 0.023 | 0.46 |

*Note:* The measurement units have been rescaled. The number of prescriptions were analysed in units of 50 prescriptions; the total duration of use and the duration of the first prescription period were expressed in years. Model 1 was adjusted for age and sex; Model 2 was adjusted for age, sex, body mass index, morbidity count, household income, highest qualification and cohabitation status. Rx = prescription; CI = confidence interval.

Sensitivity analysis: minimum duration 4.5 years

**Table S2.** Associations between the duration of lithium use and biological ageing markers, excluding individuals with a total duration of lithium use < 4.5 years

| | Model 1 (adj. for age and sex) | | | | | Model 2 (full adjustment) | | | | |
|---|---|---|---|---|---|---|---|---|---|---|
| | *n* | Beta | 95% CI | | *p* | *n* | Beta | 95% CI | | *p* |
| **Telomere length** | | | | | | | | | | |
| Rx count | 185 | 0.045 | -0.062 | 0.151 | 0.41 | 184 | 0.032 | -0.081 | 0.144 | 0.58 |
| Total duration | 185 | 0.026 | -0.013 | 0.066 | 0.19 | 184 | 0.028 | -0.013 | 0.069 | 0.18 |
| Duration first Rx | 185 | -0.015 | -0.056 | 0.025 | 0.45 | 184 | -0.009 | -0.051 | 0.033 | 0.68 |
| **Frailty index** | | | | | | | | | | |
| Rx count | 187 | 0.011 | -0.088 | 0.110 | 0.83 | 186 | 0.040 | -0.041 | 0.120 | 0.33 |
| Total duration | 187 | -0.001 | -0.039 | 0.036 | 0.95 | 186 | 0.006 | -0.024 | 0.036 | 0.70 |
| Duration first Rx | 187 | -0.019 | -0.057 | 0.018 | 0.31 | 186 | -0.006 | -0.037 | 0.024 | 0.69 |
| **Metabolomic age** | | | | | | | | | | |
| Rx count | 44 | -0.105 | -1.047 | 0.837 | 0.82 | 44 | -0.072 | -1.154 | 1.009 | 0.89 |
| Total duration | 44 | 0.043 | -0.274 | 0.361 | 0.79 | 44 | -0.047 | -0.436 | 0.342 | 0.81 |
| Duration first Rx | 44 | 0.430 | 0.111 | 0.748 | 0.01 | 44 | 0.389 | 0.018 | 0.759 | 0.04 |
| **Pulse rate** | | | | | | | | | | |
| Rx count | 189 | -0.030 | -0.139 | 0.080 | 0.59 | 188 | -0.044 | -0.159 | 0.070 | 0.45 |
| Total duration | 189 | -0.033 | -0.074 | 0.008 | 0.11 | 188 | -0.025 | -0.067 | 0.017 | 0.25 |
| Duration first Rx | 189 | 0.000 | -0.042 | 0.042 | 0.99 | 188 | -0.014 | -0.058 | 0.029 | 0.52 |

*Note:* The measurement units have been rescaled. The number of prescriptions were analysed in units of 50 prescriptions; the total duration of use and the duration of the first prescription period were expressed in years. Model 1 was adjusted for age and sex; Model 2 was adjusted for age, sex, body mass index, morbidity count, household income, highest qualification and cohabitation status. Rx = prescription; CI = confidence interval.

Sensitivity analysis: stratified by antidepressant use

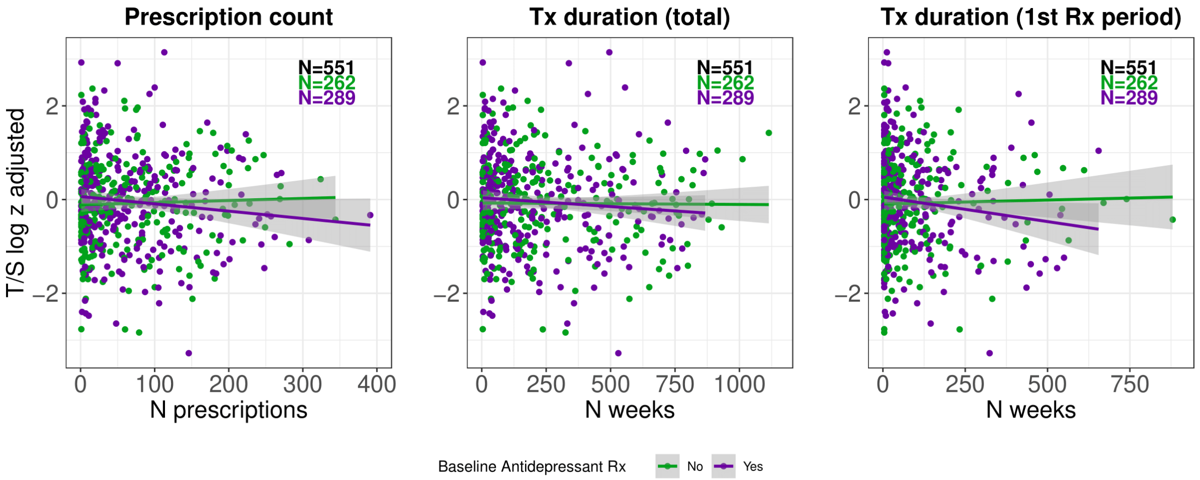

**Figure S4**. Scatter plots of telomere length (T/S ratio $\log_e$ transformed and Z standardised) by lithium prescription count, total duration of use, and duration of the first prescription period. The lines were estimated using ordinary least squares regression, and the shaded areas correspond to 95% confidence intervals. Antidepressant use (yes/no) at the time of the baseline assessment when telomere length was measured is shown in different colours. Individuals without baseline data on current antidepressant use are not shown. Tx = treatment; Rx = prescription.

Sensitivity analysis: additional adjustment for antidepressant use

**Table S3.** Associations between the duration of lithium use and biological ageing markers with additional adjustment for antidepressant use

| | Model 1 (adj. for age and sex) | | | | | Model 2 (full adjustment) | | | | |
|---|---|---|---|---|---|---|---|---|---|---|
| | *n* | Beta | 95% CI | | *p* | *n* | Beta | 95% CI | | *p* |
| **Telomere length** | | | | | | | | | | |
| Rx count | 551 | -0.017 | -0.076 | 0.042 | 0.58 | 549 | -0.014 | -0.073 | 0.046 | 0.65 |
| Total duration | 551 | -0.005 | -0.023 | 0.013 | 0.58 | 549 | -0.004 | -0.022 | 0.014 | 0.67 |
| Duration first Rx | 551 | -0.017 | -0.050 | 0.017 | 0.33 | 549 | -0.013 | -0.046 | 0.021 | 0.46 |
| **Frailty index** | | | | | | | | | | |
| Rx count | 552 | -0.046 | -0.104 | 0.012 | 0.12 | 550 | -0.044 | -0.092 | 0.004 | 0.07 |
| Total duration | 552 | -0.008 | -0.026 | 0.010 | 0.40 | 550 | -0.011 | -0.026 | 0.004 | 0.13 |
| Duration first Rx | 552 | -0.027 | -0.060 | 0.006 | 0.11 | 550 | -0.023 | -0.050 | 0.004 | 0.09 |
| **Metabolomic age** | | | | | | | | | | |
| Rx count | 115 | -0.124 | -0.696 | 0.448 | 0.67 | 115 | -0.140 | -0.707 | 0.427 | 0.62 |
| Total duration | 115 | 0.035 | -0.131 | 0.201 | 0.68 | 115 | -0.022 | -0.192 | 0.148 | 0.80 |
| Duration first Rx | 115 | 0.197 | -0.157 | 0.550 | 0.27 | 115 | 0.124 | -0.225 | 0.472 | 0.48 |
| **Pulse rate** | | | | | | | | | | |
| Rx count | 556 | -0.069 | -0.128 | -0.010 | 0.02 | 554 | -0.066 | -0.125 | -0.008 | 0.03 |
| Total duration | 556 | -0.023 | -0.041 | -0.005 | 0.01 | 554 | -0.022 | -0.040 | -0.004 | 0.02 |
| Duration first Rx | 556 | -0.019 | -0.052 | 0.015 | 0.28 | 554 | -0.020 | -0.054 | 0.013 | 0.23 |

*Note:* The measurement units have been rescaled. The number of prescriptions were analysed in units of 50 prescriptions; the total duration of use and the duration of the first prescription period were expressed in years. Model 1 was adjusted for age, sex and current antidepressant use at the time of the baseline assessment (yes/no); Model 2 was adjusted for age, current antidepressant use, sex, body mass index, morbidity count, household income, highest qualification and cohabitation status. Rx = prescription; CI = confidence interval.
